# SARS-CoV-2 Mortality Surveillance among Community Deaths brought to University Teaching Hospital Mortuary in Lusaka, Zambia, 2020

**DOI:** 10.1101/2021.11.14.21266330

**Authors:** Amos Hamukale, Jonas Z. Hines, Nyambe Sinyange, Sombo Fwoloshi, Warren Malambo, Suilanji Sivile, Stephen Chanda, Luchenga Adam Mucheleng’anga, Nkomba Kayeyi, Cordelia Maria Himwaze, Aaron Shibemba, Tally Leigh, Mazyanga L. Mazaba, Nathan Kapata, Paul Zulu, Khozya Zyambo, Francis Mupeta, Simon Agolory, Lloyd B. Mulenga, Kennedy Malama, Muzala Kapina

**Affiliations:** Zambia Field Epidemiology Training Program, Lusaka, Zambia; Centers for Disease Control and Prevention, Lusaka, Zambia; Zambia National Public Health Institute, Lusaka, Zambia; Zambia Ministry of Health, Lusaka, Zambia; Ministry of Home Affairs, Office of the State Forensic Pathologist, Nationalist Road, Lusaka, Zambia; University Teaching Hospitals, Department of Pathology and Microbiology, Nationalist Road, Lusaka, Zambia

**Keywords:** COVID-19, SARS-CoV-2, Mortality/epidemiology, Sentinel surveillance, Zambia, Africa

## Abstract

**Introduction:** During March-December 2020, Zambia recorded 20,725 confirmed COVID-19 cases, with the first wave peaking between July and August. Of the 388 COVID-19-related deaths occurring nationwide, most occurred in the community. We report findings from COVID-19 mortality surveillance among community deaths brought to the University Teaching Hospital (UTH) mortuary in Lusaka.

**Methods:** In Zambia, when a person dies in the community, and is brought into a health facility mortuary, they are recorded as ‘brought in dead’ (BID). The UTH mortuary accepts persons BID for Lusaka District, the most populated district in Zambia. We analyzed data for persons BID at UTH during 2020. We analyzed two data sources: weekly SARS-CoV-2 test results for persons BID and monthly all-cause mortality numbers among persons BID. For all-cause mortality among persons BID, monthly deaths during 2020 that were above the upper bound of the 95% confidence interval for the historic mean (2017-2019) were considered significant. Spearman’s rank test was used to correlate the overall percent positivity in Zambia with all-cause mortality and SARS-CoV-2 testing among persons BID at UTH mortuary.

**Results:** During 2020, 7,756 persons were BID at UTH (monthly range 556-810). SARS-CoV-2 testing began in April 2020, and through December 3,131 (51.9%) of 6,022 persons BID were tested. Of these, 212 (6.8%) were SARS-CoV-2 positive with weekly percent test positivity ranging from 0-32%, with the highest positivity occurring during July 2020. There were 1,139 excess persons BID from all causes at UTH mortuary in 2020 compared to the 2017-2019 mean. The monthly number of persons BID from all causes was above the upper bound of the 95% confidence interval during June-September and December.

**Conclusion:** Increases in all-cause mortality and SARS-CoV-2 test positivity among persons BID at UTH mortuary corresponded with the first peak of the COVID-19 epidemic in June and August 2020, indicating possible increased mortality related to the COVID-19 epidemic in Zambia. Combining all-cause mortality and SARS-CoV-2 testing for persons BID provides useful information about the severity of the epidemic in Lusaka and should be implemented throughout Zambia.

## Introduction

Zambia is experiencing an epidemic of severe acute respiratory syndrome coronavirus 2 (SARS-CoV-2), the causative pathogen of coronavirus disease 2019 (COVID-19). The country recorded its first two cases of COVID-19 on March 20, 2020 and its first Covid-19 related mortality on April 2, 2020. From March to December, 2020, there were 20,725 confirmed SARS-CoV-2 infections and 388 deaths attributed to Covid-19 (1). Lusaka, as the nation’s capital with over 2.7 million individuals reported the largest number of cases and deaths. Traditionally, approximately 50% of deaths in Zambia occur in the community (2).

Monitoring COVID-19 mortality provides information on the severity and impact of the COVID-19 pandemic. Deaths among persons with confirmed COVID-19 likely reflect mortality related to the SARS-CoV-2 as an underlying or associated condition but do not capture the full extent of the COVID-19 burden, whereas, deaths from all-causes can be used to estimate excess mortality and provide a more complete picture of the fatal impact of COVID-19 (3–5). Therefore, WHO recommends surveillance of confirmed COVID-19 mortality and all-cause mortality (6).

Limited information is available on COVID-19 mortality surveillance in Africa (7). A study conducted at University Teaching Hospital (UTH) in Lusaka, Zambia, found 16% SARS-CoV-2 positivity among a small sample of community and facility deaths from June to September 2020, a period which encompassed the first wave of COVID-19 in Zambia (8). In South Africa, all-cause mortality exceeded the expected deaths during peaks of COVID-19 cases in 2020-2021, with nearly 240,000 excess deaths from natural causes since March 2020 (9,10). Additionally, in South Africa, death rates were higher among males, older persons, and persons with comorbidities like cardiac disease, diabetes, hypertension, and HIV (11,12). Additionally, higher mortality was observed among hospitalized persons in South Africa during wave 2 compared to wave 1, which was attributed older age of persons admitted to the hospital, overwhelmed health care services, and the beta SARS-CoV-2 variant (13).

In April 2020, the Zambia Ministry of Health (MOH) began testing all persons BID with a history of respiratory symptoms proximal to death for SARS-CoV-2 as part of its COVID-19 surveillance strategy (14). As cases peaked during the first wave, beginning in May through August 2020, this strategy was expanded to include all persons BID. Although BID testing was intended to be done throughout Zambia, in practice, it was variably implemented with Lusaka District being the most consistent. We report on the implementation of COVID-19 mortality surveillance by the MOH at UTH mortuary through testing of SARS-CoV-2 and all-cause mortality among persons BID.

## Methods

### Study Design

We analyzed data on persons BID at UTH mortuary during January-December 2020 to assess whether SARS-CoV-2 testing of persons BID and/or all-cause mortality provided a measure of the epidemic’s impact in Zambia. In Zambia, about half of deaths occur in the community (15). When a deceased person is brought to a health facility mortuary, they are registered as ‘brought in dead’ (BID). Persons who die within 48 hours of admission to the health facility are also sometimes counted as BID in Zambia. However, all persons included in this analysis had died prior to arrival at UTH mortuary. In Lusaka District, registering a death requires a death certificate issued by a medical officer or pathologist affiliated with a hospital mortuary; thus approximately 90% of community deaths are registered in Lusaka District in contrast with 20% in the rest of Zambia (16). The UTH mortuary is the largest mortuary in the nation, with an approximate 500 body storage capacity and it accepts BID every day and at all hours for Lusaka District. During 2020, over 80% of all SARS-CoV-2 testing results among persons BID in Zambia were reported from Lusaka District (Zambia National Public Health Institute, unpublished data).

### Data Collection

Zambia implemented BID testing for SARS-CoV-2 in April 2020 (14). The Zambia National Public Health Institute (ZNPHI) maintains a database of all SARS-CoV-2 testing results that includes information for testing location and case identification strategies including contact tracing, points-of-entry (i.e., at air and land borders), and person BID. We obtained the SARS-CoV-2 test results for the persons BID at UTH mortuary from ZNPHI. Data on age, sex, date of death, and date of testing were abstracted from the ZNPHI database for the analysis. ZNPHI extracted data were externally validated with the UTH mortuary registers to verify representativeness and completeness of information. SARS-CoV-2 testing capacity was limited in Zambia during 2020 and not all persons BID were tested for SARS-CoV-2 at UTH mortuary. No preference was given of who to test at UTH mortuary.

Trained mortuary attendants performed specimen collection for SARS-CoV-2 testing within 24 hours of receipt of the body. A nasopharyngeal sample was collected using scored swabs inserted into a deceased person’s nasopharynx using standard personal protective equipment and measures during specimen collection. The specimen was then transported in viral transport media on ice to the virology lab at UTH where it was tested using the real-time polymerase chain reaction test to detect SARS-CoV-2 RNA. Rapid antigen tests were not used for testing persons BID during this study period.

All-cause mortality is defined as the occurrence of death regardless of the underlying cause-of-death. For all-cause mortality data, Zambia MOH routinely collects aggregate-level metrics from health facilities—including persons who died in the facility or were BID—on a monthly basis and stores it in the District Health Information System 2 (DHIS-2) (17). The monthly numbers of persons BID at UTH from 2017 through 2020 were obtained from DHIS-2 to compare the 2020 trend against the historic mean from 2017-2019. DHIS-2 data was cross-referenced with facility-level registers when available, noting some documents were not identified for every month from the 2017-2019 time period.

### Data Analysis

For confirmed SARS-CoV-2 deaths among persons BID, the percent test positivity was calculated as the number of persons BID testing SARS-CoV-2 positive divided by the total number of persons BID who were tested for SARS-CoV-2 infection. The weekly positivity trend among persons BID at UTH was compared to the weekly trend in test positivity for all SARS-CoV-2 tests in Zambia. Additionally, results were disaggregated by age and sex of decedents. To measure the association between demographic variables and SARS-CoV-2 testing, we used Chi-square for categorical variables, sex and age group, and Wilcoxon rank sum test for age. The strength of correlation between SARS-CoV-2 positivity among persons BID, and overall SARS-CoV-2 testing positivity, was measured using Spearman’s rank correlation coefficient. There were 17 (<1.0%) observations excluded from the April to December 2020 analysis due to missing data for age and sex.

To estimate excess mortality from COVID-19, the monthly number of persons BID from all causes at UTH in 2020 was compared to its historic trends for 2017-2019. Excess mortality for a given month was defined as the difference between the 2020 value and the historic mean from 2017-2019. The difference was considered significant if it deviated beyond the upper bound of the 95% confidence interval, calculated by using the sample standard deviations of the historic mean number of deaths among persons BID from 2017-2019 (18). An excel-based tool was used to plot the 2020 data and visually compare the trend against the mean and 95% confidence interval of the historic data from 2017-2019 (19). Additionally, the strength of association between all-cause mortality among persons BID and overall SARS-CoV-2 test positivity in Zambia was measured through Spearman’s rank correlation coefficient.

Data distribution of continuous variables was checked for normality using Shapiro-Wilk W-test. For non-parametric variables median and interquartile range was reported while mean and standard deviation was reported for parametric variables. Analyses were performed using STATA version 14.0 (Stata Corp., College Station, TX, USA) and Microsoft Excel 2016 (Microsoft, Redmond, WA, USA) software. This project was reviewed by the Zambia National Health Research Authority and exempted from ethical review. The activity was reviewed by the US Centers for Disease Control and Prevention (CDC) and conducted consistent with applicable US federal law and CDC policy.

## Results

From January to December 2020, 7,756 persons were BID for all causes at UTH mortuary. During January-March 2020, no SARS-CoV-2 testing occurred at UTH mortuary. Between April to December 2020, 6,022 persons were BID and among these, males were significantly younger than females (median age: 41 years versus 48 years; p<0.01). Of the 6022 BIDs 3,131 (51.9%) were tested for SARS-CoV-2 (Table 1). Persons BID who were tested for SARS-CoV-2 were significantly older than those who were not tested (median age: 45 years versus 42 years; p<0.01). There was no difference in proportion tested by sex (p=0.424).

**TABLE 1:**
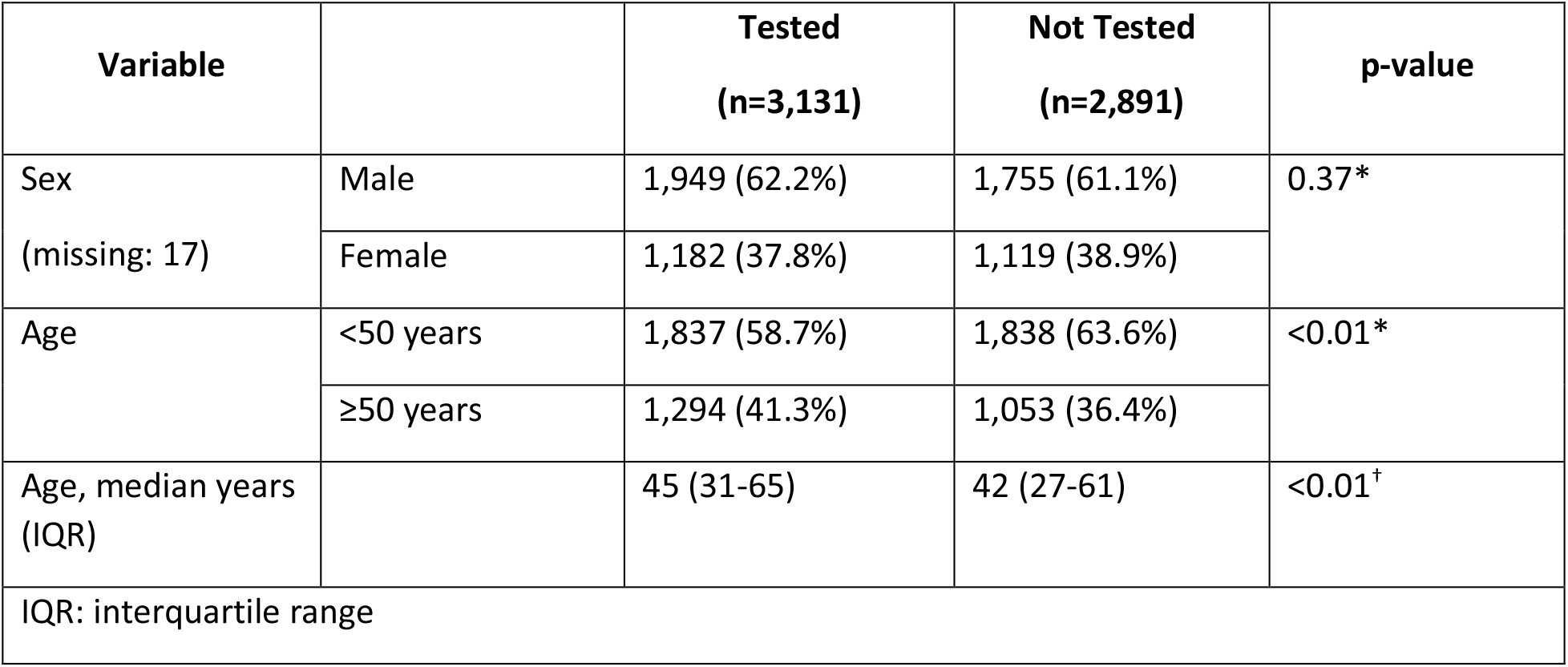
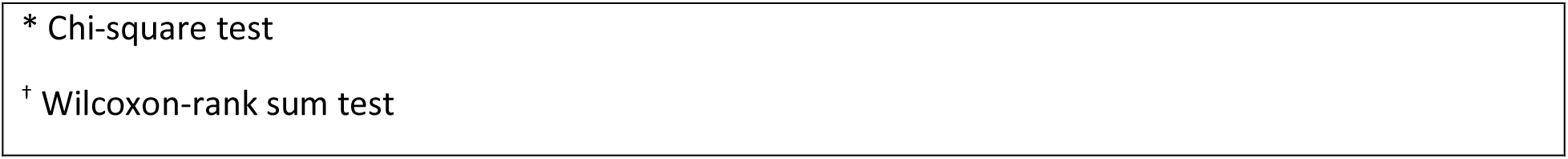
Characteristics of persons brought-in-dead based on SARS-CoV-2 testing status — University Teaching Hospital, Lusaka, Zambia, April-December 2020 (N=6,022)

| Variable |  | Tested<br>(n=3,131) | Not Tested<br>(n=2,891) | p-value |
| --- | --- | --- | --- | --- |
| Sex<br>(missing: 17) | Male | 1,949 (62.2%) | 1,755 (61.1%) | 0.37* |
|  | Female | 1,182 (37.8%) | 1,119 (38.9%) |  |
| Age | <50 years | 1,837 (58.7%) | 1,838 (63.6%) | <0.01* |
|  | ≥50 years | 1,294 (41.3%) | 1,053 (36.4%) |  |
| Age, median years<br>(IQR) |  | 45 (31-65) | 42 (27-61) | <0.01<sup>†</sup> |
| IQR: interquartile range |  |  |  |  |

\* Chi-square test
† Wilcoxon-rank sum test

Of the 3,131 persons BID to UTH mortuary who were tested for SARS-CoV-2 from April to December 2020, 212 (6.8%) tested positive for SARS-CoV-2 infection (Table 2). There was no difference in test positivity between males and females (p=0.47). Persons BID who tested positive for SARS-CoV-2 were significantly older than those testing negative (median age: 50 versus 42 years; p<0.01).

**TABLE 2:** Characteristics of persons brought-in-dead by SARS-CoV-2 test result — University Teaching Hospital, Lusaka, Zambia, April-December 2020 (N= 3,131)

| Variable |  | SARS-CoV-2 positive<br>(n= 212) | SARS-CoV-2 negative<br>(n=2,919) | p-value |
| --- | --- | --- | --- | --- |
| Sex, n (%) | Male | 127 (6.5%) | 1822 (93.5%) | 0.47* |
|  | Female | 85 (7.2%) | 1097 (92.8%) |  |
| Age, n (%) | <50 years | 99 (5.4%) | 1738 (94.6%) | <0.01* |
|  | ≥50 years | 113 (8.7%) | 1181 (91.3%) |  |
| Age, median years (IQR) |  | 50 (34-68) | 42 (27-60) | <0.01<sup>†</sup> |
| IQR: interquartile range |  |  |  |  |
| * Chi-square test |  |  |  |  |
| † Wilcoxon-rank sum test |  |  |  |  |

During 2020, there were 20,725 (3.4%) positive SARS-CoV-2 results out of 601,003 tests in Zambia. Beginning in mid-June through July 2020, Lusaka experienced a surge in the number of SARS-CoV-2 persons BIDs testing positive for SARS-CoV-2, from a mean of 1.5 cases per week during April to mid-June 2020, to 24 cases per week during mid-June to July (Figure 1). SARS-CoV-2 positivity among persons BID at UTH mortuary was correlated with overall SARS-CoV-2 test positivity in Zambia (rho=0.5, p<0.01).

**Figure 1:**
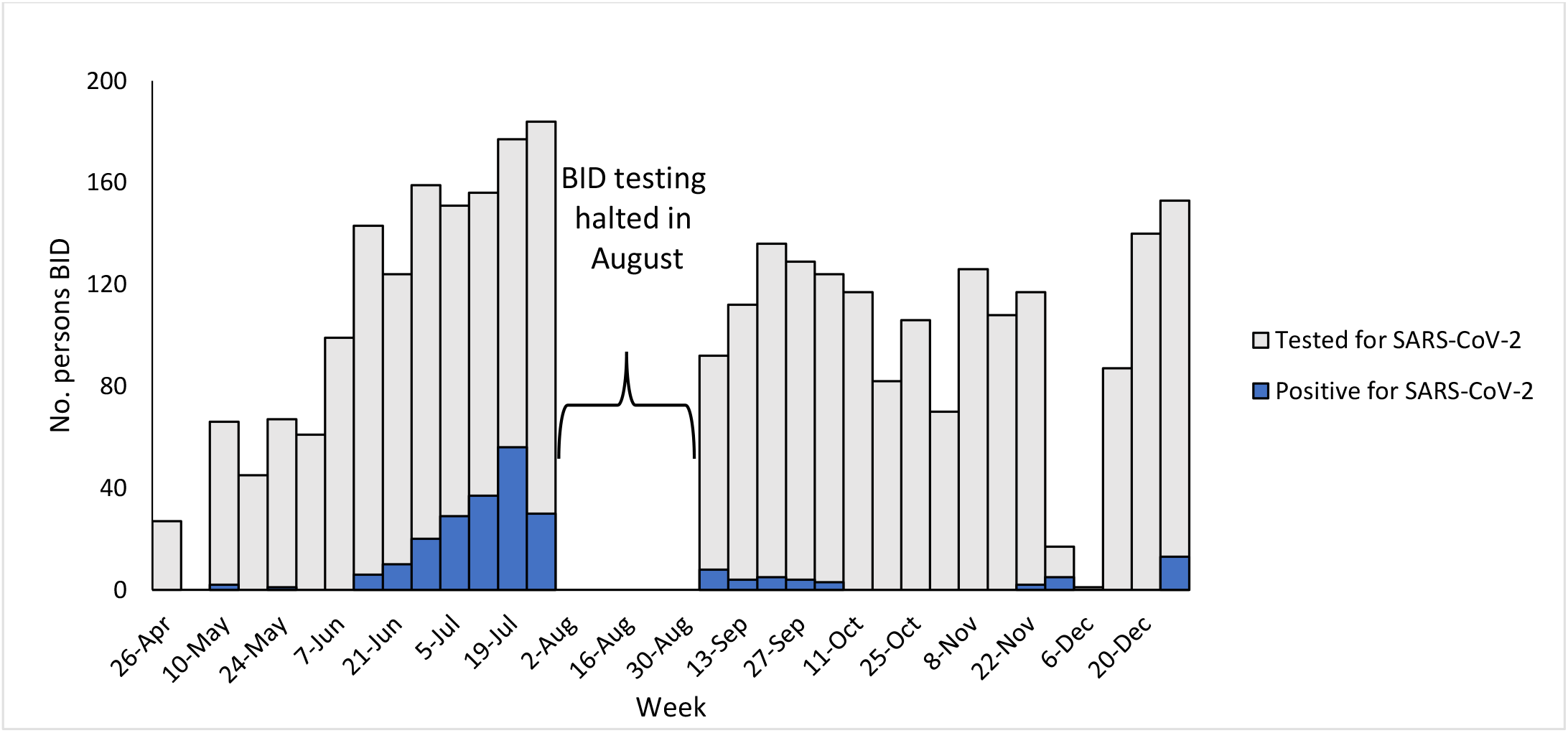
Weekly SARS-CoV-2 test results for persons brought-in-dead — University Teaching Hospital, Lusaka, Zambia, April-December 2020 (N= 3,131)^*^ **\***.No SARS-CoV-2 testing was performed at UTH mortuary during January to March 2020.

**Figure 2:**
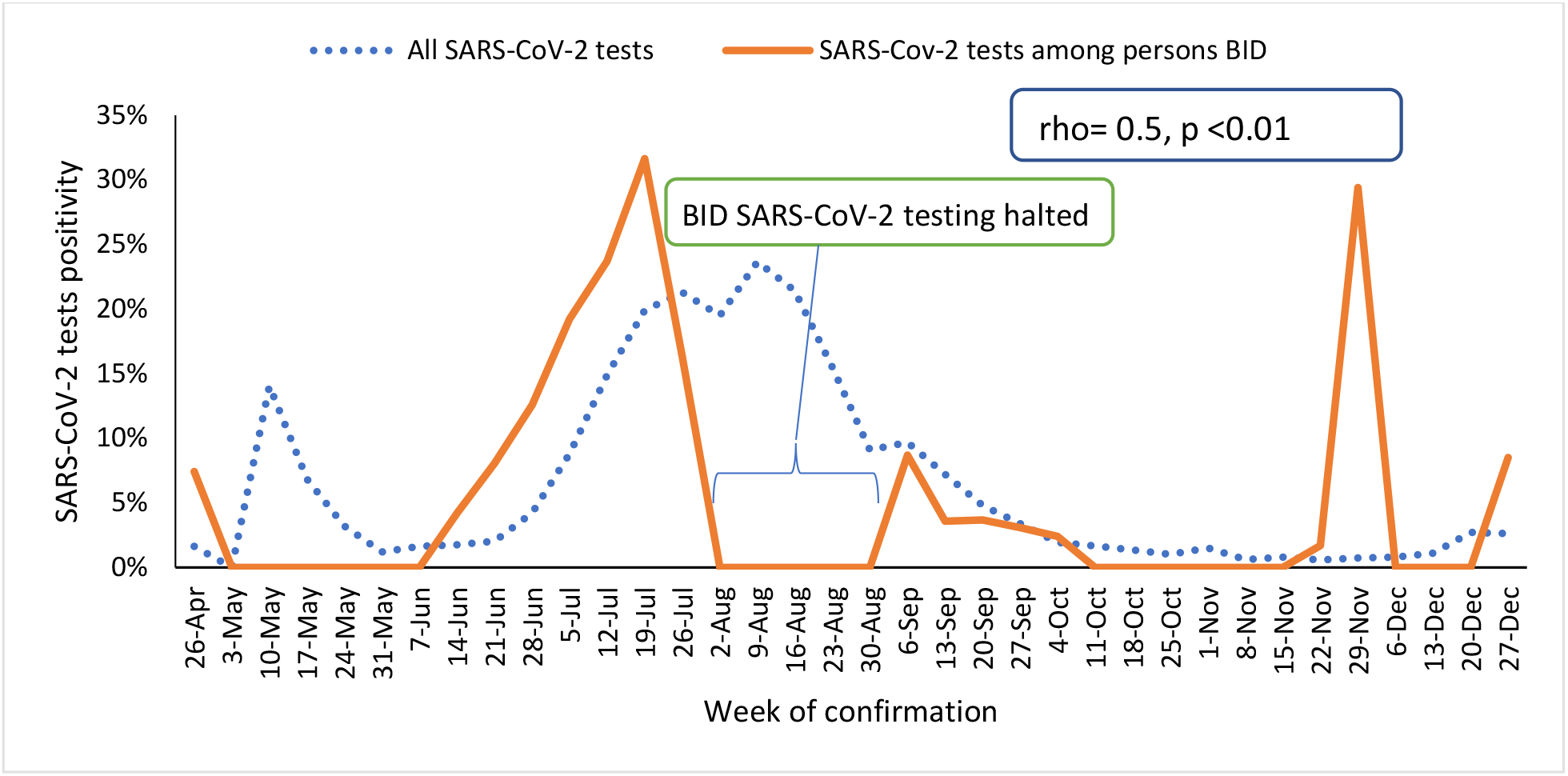
SARS-CoV-2 test positivity among persons brought-in-dead to University Teaching Hospital Mortuary in Lusaka and all persons tested for SARS-CoV-2 in Zambia, April - December 2020 (n= 3131)

For the period January to December 2020, the mean number of BIDs per month at UTH mortuary was 646 (SD=84.8) compared to a mean number of 551 (SD=29.8) per month for 2017-2019 (p<0.01). There were 1,139 excess deaths from all causes at UTH mortuary during 2020 compared to the 2017-2019 historic mean. During June, July, August, September, November and December 2020, all-cause mortality among persons BID exceeded the upper bound of the 95% confidence interval of the 2017-2019 historic mean (Figure 3). The peaks in all-cause mortality at UTH mortuary corresponded to peaks in the overall case counts in Zambia. There was a strong positive correlation observed between persons BID for all causes at UTH mortuary and overall SARS-CoV-2 test positivity in Zambia (rho=0.80, p=0.01).

**Figure 3:**
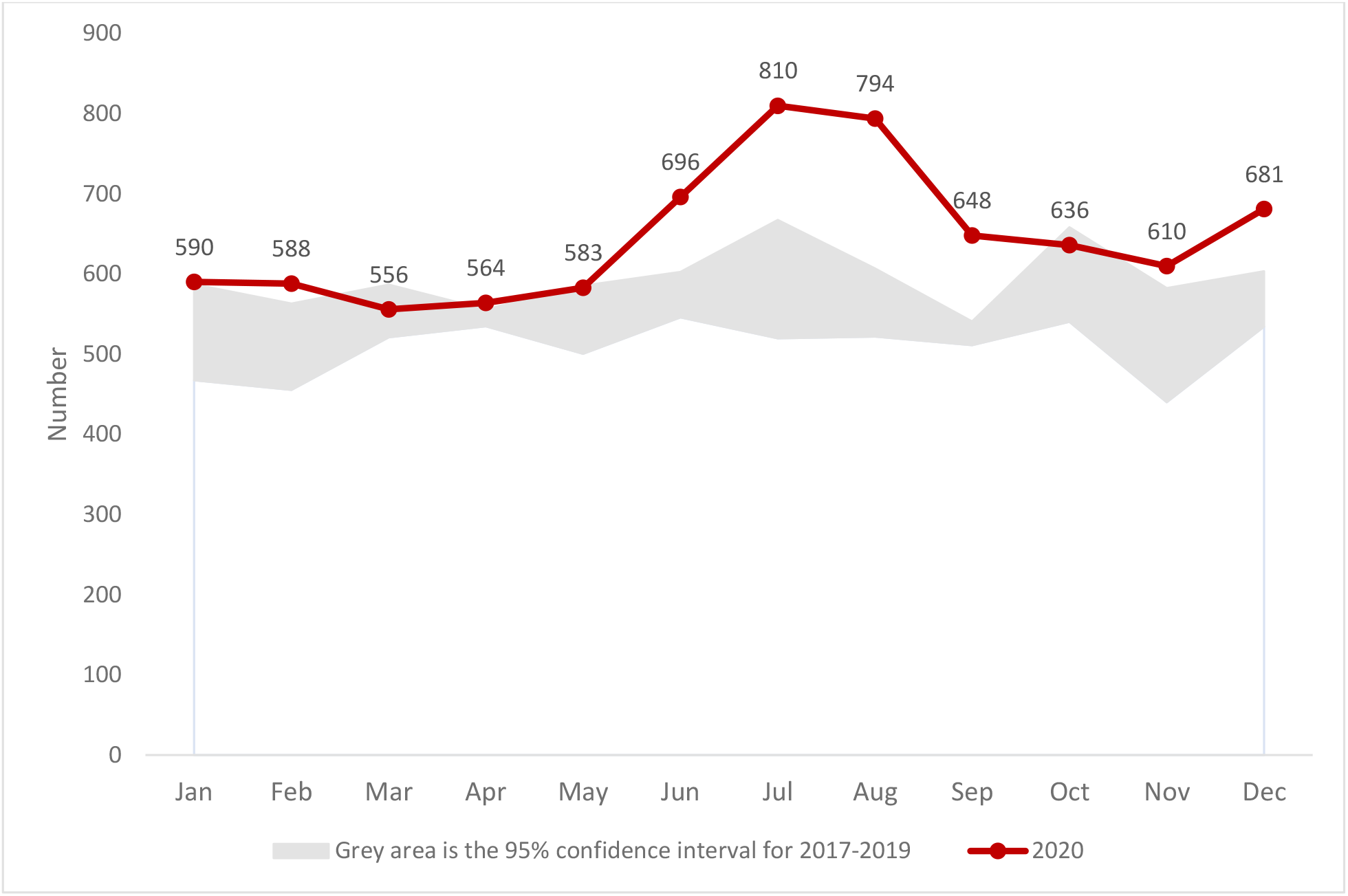
Time trend of all-cause mortality among persons brought-in-dead in 2020 compared to the historic trend from 2017-2019 — University Teaching Hospital, Lusaka, Zambia (n=7,756)

## Discussion

During the 2020 COVID-19 epidemic in Zambia, the surveillance of persons BID at UTH mortuary, through testing for SARS-CoV-2 and all-cause mortality analysis, correlated with the epidemiologic situation in the country. Peaks in SARS-CoV-2 test positivity among persons BID and excess all-cause mortality at UTH corresponded to both peaks in overall SARS-CoV-2 test positivity (i.e., periods of community transmission) in Zambia. Persons BID to UTH mortuary had a high SARS-CoV-2 test weekly positivity in 2020 at UTH mortuary compared with weekly positivity of all SARS-CoV-2 tests in 2020. Furthermore, the high prevalence of SARS-CoV-2 positivity found among persons BID at UTH mortuary suggest more people might have died from COVID-19 than official statistics capture (8). This is not surprising considering the low case detection proportion of SARS-CoV-2 infections observed the first wave in Zambia (20). Over two-thirds of Covid-19 deaths were identified via test persons BID. Mortality surveillance at UTH mortuary provided additional information about the severity of the COVID-19 epidemic in Lusaka. A more complete picture of the toll of the COVID-19 epidemic could be gathered if this approach was systematically expanded throughout Zambia.

A person BID testing positive for SARS-CoV-2 is a sentinel event signaling undetected cases in the community. It has been difficult to determine the extent of SARS-CoV-2 spread within the general population in Zambia because testing capacity was generally been below recommend levels (21,22) in 2020 and most infections were asymptomatic (20). In the absence of adequate testing resources, testing persons BID might help estimate the severity and extent of spread of the COVID-19 epidemic in the population. Moreover, testing persons BID might serve as a case identification strategy by linking to cases and clusters among contacts. For instance, a SARS-CoV-2 positivity among persons BID was approximately twice that of the general population in2020.

Excess all-cause mortality among persons BID was correlated with increased overall SARS-CoV-2 positivity in Zambia. This implies that analysis of excess all-cause mortality among persons BID provides a proxy indicator of SARS-CoV-2 cases in the population during the Covid-19 pandemic. In South Africa, all-cause mortality surveillance has been a useful surveillance strategy during the COVID-19 epidemic (9,10). Combining BID testing with excess mortality analysis among persons BID resulted in a robust sentinel surveillance system in Lusaka that helped demonstrate a more accurate toll of the SARS-CoV-2 epidemic in Zambia, including estimating excess deaths that might have been related to COVID-19 in the absence of BID testing, as occurred at UTH mortuary in August 2020.

Tracking SARS-CoV-2 among persons BID might be a useful indicator of access to care. If the proportion of persons dying from COVID-19 in the community is very high, risk communication interventions might be directed toward such areas where cases resided to bolster health seeking behavior for COVID-19. Ideally, the public is aware of the signs and symptoms of COVID-19 and has information on when to seek care for COVID-19. However, high numbers of persons BID testing positive for SARS-CoV-2 could signal gaps in effective public health risk communications. Moreover, utilizing the data provided by BID testing might be an effective risk communication strategy; if the public is aware of the severity and impact of SARS-CoV-2 on the community, it might be more willing to follow preventive measures proven to decrease SARS-CoV-2 transmission like mask wearing and social distancing.

SARS-CoV-2 disproportionately affected older persons at UTH mortuary. Older age is an established risk factor for severe COVID-19 disease and death (23–25). This finding could be attributed to higher prevalence of underlying co-morbidities in older persons (which was not measured in this study) versus a greater proportion of unnatural deaths (which are less likely to be Covid-19-related) among younger persons BID at UTH mortuary. The age and sex findings in this study are similar to what was found in the handful of studies on COVID-19 mortality in Africa (11,26). Health care teams should intensify COVID-19 screening and management of older persons presenting with underlying comorbidities.

The findings in this study were similar to a small study of COVID-19 mortality at UTH mortuary (8), which was expected given the studies drew from the same population. However, our study was larger and included all persons BID with SARS-CoV-2 tests performed as part of the MOH’s surveillance strategy in Zambia (8). Our study relied upon routinely collected data, which may be a more sustainable surveillance approach than was deployed by Mwananyanda et al. However, this also meant we had less detailed individual-level data. Data from verbal autopsies at UTH mortuary could be linked with testing data from persons BID to improve Covid-19 mortality surveillance in Zambia and elsewhere.

This study was subject to several limitations. We utilized data that were available from routine SARS-CoV-2 testing of persons BID and many persons BID were not tested for SARS-CoV-2, including no one during the month of August 2020 which was during the first peak of the epidemic in Zambia. During August 2020, case counts, and percent test positivity remained elevated, but when BID testing resumed in September 2020, test positivity had substantially reduced. Next, excess all-cause mortality does not necessarily reflect the impact of COVID-19, although there was an absence of other large-scale health events in Lusaka that could explain the peaks observed in the data. However, it is possible that patients with other underlying health conditions did not access health services because of fear of COVID-19 and some of them may have died as a result. Although the UTH mortuary is the largest in Zambia, and the majority of SARS-CoV-2 testing among persons BID have been reported from this facility, the findings from this study might not be representative of other mortuaries in Zambia or Africa. Lastly, as an observational study, associations reported in this study do not necessarily reflect a causal relationship.

Mortality surveillance at UTH mortuary in Lusaka correlated with SARS-CoV-2 transmission in the general population. Mortality surveillance should be implemented to ensure timely detection and management of COVID-19 cases in resource-limited settings where testing rates are low and more likely to underestimate the extent and impact the disease on the community. Putting the data into context with other specific causes of death in Zambia is difficult given the likely under-capture of COVID-19 deaths in Zambia. Ultimately, advancing mortality surveillance in resource-limited countries involves instituting accurate and standard completion of the death certificate for facility and community death, including ascertaining underlying cause of death, for use in mortality surveillance. However, in the absence of a high coverage civil registration vital statistics system (27), strategies like all-cause mortality analyses and testing deceased persons for SARS-CoV-2, especially if coupled with verbal autopsies, are potentially useful surveillance strategies.

## Data Availability

All data produced in the present study are available upon reasonable request to the authors

## Authorship Disclaimer

*The findings and conclusions in this report are those of the author(s) and do not necessarily represent the official position of the funding agencies*.

## References

1. Zambia National Public Health Institute. Zambia COVID-19 Statistics; Daily Status Update [Internet]. Available from: https://www.facebook.com/ZMPublicHealth/

2. Central Statistical Office. Zambia SAVVY repoty 2015-16: mortality and causes of death information from verbal autopsy [Internet]. 2018. Available from: https://www.zamstats.gov.zm/phocadownload/Demography/SAVVYReport2015-16.pdf

3. Rossen LM, Branum AM, Ahmad FB, Sutton PD, Anderson RN. Notes from the Field: Update on Excess Deaths Associated with the COVID-19 Pandemic — United States, January 26, 2020–February 27, 2021. 2021;70(15):2020–1.

4. Department of Health and Mental Hygiene NYC. Preliminary Estimate of Excess Mortality During the COVID-19 Outbreak — New York City, March 11-May 2, 2020. Morb Mortal Wkly Rep. 2020;69(19):603–5.

5. Odone A, Delmonte D, Gaetti G, Signorelli C. Doubled mortality rate during the COVID-19 pandemic in Italy: quantifying what is not captured by surveillance. Public Health [Internet]. 2020; Available from: https://doi.org/10.1016/j.puhe.2020.11.016

6. World Health Organization. Public health surveillance for COVID-19 [Internet]. 2020. Available from: https://www.who.int/publications/i/item/who-2019-nCoV-surveillanceguidance-2020.7

7. Shveda K, Cardoso K, Adamou L, Martens J, Ericsson N. Measuring Africa’s Data Gap: The cost of not counting the dead. BBC [Internet]. 2021; Available from: https://www.bbc.com/news/world-africa-55674139

8. Mwananyanda L, Gill CJ, MacLeod W, Kwenda G, Pieciak R, Mupila Z, et al. Covid-19 deaths in Africa: prospective systematic postmortem surveillance study. BMJ. 2021;372.

9. South African Medical Research Council. Report on Weekly Deaths in South Africa [Internet]. 2021. Available from: https://www.samrc.ac.za/reports/report-weekly-deaths-south-africa

10. Bradshaw D, Laubscher R, Dorrington R, Groenewald P, Moultrie T. Report on Weekly Deaths in South Africa; 8-14 August 2021 (Week 32) [Internet]. Available from: https://www.samrc.ac.za/sites/default/files/files/2021-08-18/weekly14Aug2021.pdf

11. Pillay-Van Wyk V, Bradshaw D, Groenewald P, Seocharan I, Manda S, Roomaney RA, et al. COVID-19 deaths in South Africa: 99 days since South Africa’s first death. South African Med J. 2020;110(11):1093–9.

12. Jassat W, Cohen C, Tempia S, Masha M, Goldstein S, Kufa T, et al. Articles Risk factors for COVID-19-related in-hospital mortality in a high HIV and tuberculosis prevalence setting in South Africa: a cohort study. 2021;3018(21):1–14.

13. Jassat W, Mudara C, Ozougwu L, Tempia S, Blumberg L, Davies M, et al. Difference in mortality among individuals admitted to hospital with COVID-19 during the first and second waves in South Africa: a cohort study. Lancet Glob Heal. 2021;S2214-109X(21)00289-8.

14. Zambia Ministry of Health. Implementation Strategy – Active Screening for COVID-19 in Health Facilities. Lusaka; 2020.

15. Mapoma CC, Munkombwe B, Mwango C, Bwalya BB, Kalindi A, Gona NP. Application of verbal autopsy in routine civil registration in Lusaka District of Zambia. BMC Health Serv Res. 2021;21(1):1–11.

16. Central Statistical Office and Ministry of Home Affairs. 2019 Vital Statistics Report. Lusaka, Zambia; 2021.

17. DHIS2 In Action [Internet]. Available from: https://dhis2.org/in-action/

18. Vital Strategies. Revealing the Toll of COVID-19: A Technical Package for Rapid Mortality Surveillance and Epidemic Response [Internet]. 2020. Available from: https://www.who.int/publications/i/item/revealing-the-toll-of-covid-19

19. Vital Strategies, Prevent Epidemics. Estimating Excess Mortality from Covid-19 [Internet]. 2020. Available from: https://vital.ent.box.com/v/ExcessMortalityCalculator

20. Mulenga LB, Hines JZ, Fwoloshi S, Chirwa L, Siwingwa M, Yingst S, et al. Prevalence of SARS-CoV-2 in six districts in Zambia in July, 2020: a cross-sectional cluster sample survey. Lancet Glob Heal. 2021;(21):1–9.

21. Africa Centres for Disease Control and Prevention. Monitoring and Evaluation Framework for the Partnership to Accelerate COVID-19 Testing [Internet]. Addis Ababa; 2020. Available from: https://africacdc.org/download/monitoring-and-evaluation-framework-for-the-partnership-to-accelerate-covid-19-testing/

22. Vital Strategies. Annex 1: How to create a COVID-19 alert-level system and supporting communication tools in resource-constrained settings [Internet]. 2020. Available from: https://preventepidemics.org/wp-content/uploads/2020/05/Annex-1_COVID-19-alert-level-system-in-resource-constrained-settings_FINAL.pdf

23. Rosenthal N, Cao Z, Gundrum J, Sianis J, Safo S. Risk Factors Associated With In-Hospital Mortality in a US National Sample of Patients With COVID-19. JAMA Netw open. 2020;3(12):e2029058.

24. Bialek S, Boundy E, Bowen V, Chow N, Cohn A, Dowling N, et al. Severe Outcomes Among Patients with Coronavirus Disease 2019 (COVID-19) — United States, February 12–March 16, 2020. MMWR Morb Mortal Wkly Rep. 2020;69(12).

25. Williamson EJ, Walker AJ, Bhaskaran K, Bacon S, Bates C, Morton CE, et al. Factors associated with COVID-19-related death using OpenSAFELY. Nature. 2020;584(7821).

26. Boulle AA, Davies M, Hussey H, Morden E, Vundle Z, Zweigenthal V, et al. Risk factors for COVID-19 death in a population cohort study from the Western Cape Province, South Africa.: 1–31.

27. Gundlapalli A V., Lavery AM, Boehmer TK, Beach MJ, Walke HT, Sutton PD, et al. Death Certificate–Based ICD-10 Diagnosis Codes for COVID-19 Mortality Surveillance — United States, January–December 2020. MMWR Morb Mortal Wkly Rep. 2021;70(14):523–7.

